# The Relationship of Duffy Gene Polymorphism, High Sensitivity C-Reactive Protein, and Long-term Outcomes

**DOI:** 10.1101/2023.08.03.23293626

**Authors:** Edward T. Ha, Kent D. Taylor, Laura M Raffield, Matt Briggs, Aaron Yee, Olivier Elemento, Manish Parikh, Stephen J. Peterson, William Frishman, Robert E. Gerszten, James G. Wilson, Karl Kelsey, Usman A. Tahir, Alex Reiner, Paul Auer, Teresa Seeman, Stephen S. Rich, April P. Carson, Wendy S. Post, Jerome I. Rotter, Wilbert S. Aronow

## Abstract

**Background:** Black adults have higher incidence of all-cause death and worse cardiovascular outcomes when compared to other populations. The Duffy chemokine receptor is not expressed in a large majority of Black adults and the clinical implications of this are unclear.

**Methods:** Here, we investigated the relationship of Duffy receptor status, high-sensitivity C-reactive protein (hs-CRP), and long-term cardiovascular outcomes in Black members of two contemporary, longitudinal cohort studies (the Jackson Heart Study and Multi-Ethnic Study of Atherosclerosis). Data on 4,307 Black participants (2,942 Duffy null and 1,365 Duffy receptor positive, as defined using Single Nucleotide Polymorphism (SNP) rs2814778) were included in this analysis.

**Results:** Duffy null was not independently associated with elevated levels of serum hs-CRP levels once conditioning for known *CRP* locus alleles in linkage disequilibrium with the Duffy gene. Duffy null status was not found to be independently associated with higher incidence of all-cause mortality or secondary outcomes after adjusting for possible confounders in Black participants.

**Conclusions:** These findings suggest that increased levels of hs-CRP found in Duffy null individuals is due to co-inheritance of CRP alleles known to influence circulating levels hs-CRP and that Duffy null status was not associated with worse adverse outcomes over the follow-up period in this cohort of well-balanced Black participants.

## Introduction

In the United States, Black individuals have a particularly high burden of adverse cardiovascular outcomes, which current literature suggests is largely due to adverse social determinants of health-related exposures^1^. Black populations are understudied in human genetics and cardiovascular disease epidemiology analyses, and further study of putative risk factors is needed^2^. Most individuals of West African descent are homozygous for a single-nucleotide polymorphism (SNP) in the *DARC* gene, which defines the Duffy blood group system, leading to loss of expression of the Duffy Antigen for Chemokines (Duffy null) in these individuals. This mutation is extremely rare in other populations^3,4^. Initially thought of as a benign phenotype associated with ‘ethnic’ neutropenia, through its functioning as an atypical chemokine receptor, the Duffy receptor may modulate host response to inflammation, with potential implications in a range of human pathologies including cancer and cardiovascular disease^3,5–7^. The current literature is limited however, and few associations of the Duffy allele have been identified in the GWAS literature. Recently, it was reported that the SNP rs2814778 may be associated with differences in baseline levels of high-sensitivity C-reactive protein (hs-CRP) in MESA (n=1344, p=0.02)^8^. The purpose of this study was to investigate the association of Duffy receptor status and hs-CRP, and its effect on long-term cardiovascular outcomes, in two contemporary, longitudinal cohort studies.

## Methods

The design and protocol of the Jackson Heart Study (JHS) has been previously described^9,10^. Briefly, we analyzed follow-up data since enrollment from JHS: an ongoing, prospective study in the United States that enrolled 5,306 Black participants (age range, 20-95 years) with and without cardiovascular disease, residing in the Jackson, Mississippi metropolitan area between September 2000 and March 2004. The JHS protocol and all data collection procedures were approved by the Institutional Review Boards (IRB) of University of Mississippi Medical Center, Jackson State University, and Tougaloo College. All study participants provided written informed consent.

The design of Multi-Ethnic Study of Atherosclerosis (MESA) (ClinicalTrials.gov: NCT00005487) has been previously described^11^. Briefly, we analyzed follow-up data over 15 years from MESA: an ongoing multicenter, prospective population-based study in the United States that enrolled 6,814 participants (age range, 45-84 years) between 2000 and 2002. In MESA, approximately 38 percent of the recruited participants are White, 28 percent Black, 22 percent Hispanic, and 12 percent Asian. The MESA study was approved by the institutional review boards of each of the participating field sites in the United States (Wake Forest University, Winston-Salem, NC; Columbia University, New York City, NY; Johns Hopkins University, Baltimore, MD; University of Minnesota, Minneapolis, MD; Northwestern University, Evanston, IL; and University of California, Los Angeles, CA), and all participants provided written informed consent. All sites were compliant with the Health Insurance Portability and Accountability Act.

This specific study was approved by the IRB at New York Presbyterian-Brooklyn Methodist Hospital, a tertiary center and clinical affiliate of Weill Cornell Medicine. The current analysis included a total of 4,307 Black participants (901 from MESA and 3,406 from JHS) whose genotypes at rs2814778 were known through whole genome sequencing performed through the NHLBI Trans-Omics for Precision Medicine (TOPMed) program for MESA participants and through genotyping and subsequent imputation from the NHLBI Candidate Gene Association Resource for JHS. Whole genome sequencing data is available through restricted access via the NCBI database of Genotypes and Phenotypes (dbGaP). dbGaP accession numbers for JHS (phs000964/phs002256.v1.p1) and MESA (phs001416.v1.p1) may require IRB and dbGaP Data Access Committee (DAC) approval. Whole genome sequencing with mean read coverage of 30X was performed by TOPMed sequencing centers and is described in detail at https://www.nhlbiwgs.org/topmed-whole-genome-sequencing-methods-freeze-8. Whole genome sequencing for the JHS cohort was imputed through the 1000 Genome Project using Minimac3 on the Michigan Imputation Server^1213^. The reference panel includes 5,008 haplotypes from 26 populations across the world (http://www.internationalgenome.org). Prior to imputation, SNPs were filtered for minor allele frequency ≥1%, call rate ≥ 90%, HWE p-value > 10-6, as well as exclusion of sites with invalid or mismatched alleles for the reference panel.

Rs2814778 SNP genotype CC (null mutation) was designated as Duffy null, and rs2814778 genotype TT/CT designated as Duffy receptor positive. This variant is a fail variant in TOPMed freeze 8 based on the excess heterozygosity filter: https://bravo.sph.umich.edu/freeze8/hg38/variant/snv/1-159204893-T-C. The Duffy null genotype is highly correlated with African ancestry and any results attributed to the null genotype may be due to other variants more common in those of African ancestry inherited with the null genotype. Thus, using genetic data, each participants’ genome was compared to reference genomes of descent from the African continent versus the European continent and the “percentage” of African descent was calculated using ADMIXTURE 1.3 to control for this possibility. The reference genomes were obtained from the Human Genome Diversity Project (https://www.internationalgenome.org/data-portal/data-collection/hgdp).

All serum samples were drawn on the initial visit unless otherwise specified. In JHS, fasting blood samples were drawn and processed using a standardized protocol and sent to central laboratories at the Collaborative Studies Clinical Laboratory at Fairview University of Minnesota Medical Center (Minneapolis, MN, USA) for measurement of glucose, cholesterol and estimated glomerular filtration rate^14^. Serum glucose was measured by rate reflectance spectrophotometry using thin-film adaptation of the glucose oxidase method on a Vitros analyzer (Ortho Clinical Diagnostics, Rochester, NY, USA). Total cholesterol was measured using a cholesterol oxidase method (Roche Diagnostics, Indianapolis, IN, USA) on the Roche Cobas Fara centrifugal analyzer. Serum creatinine was measured by rate reflectance spectrophotometry using thin-film adaptation of the creatinine amidinohydrolase method on the Vitros analyzer (Ortho Clinical Diagnostics) and calibrated to Cleveland Clinic standards.

In MESA, blood was drawn after a 12 hour fast from participants, and aliquots (approximately 65 aliquots per participant) were prepared for central analysis and for storage at the University of Minnesota. Lipids were assayed on thawed EDTA plasma within 2 weeks of sample collection, using Centers for Disease Control Prevention/NHLBI standards. High-density lipid cholesterol (HDL-C) was measured using the cholesterol oxidase method (Roche Diagnostics, Indianapolis, Indiana) after precipitation of non-HDL-C with magnesium/dextran (coefficient of variation 2.9%). LDL-C was calculated using Friedewald equation^15^. Serum creatinine was measured using colorimetry with a Johnson & Johnson Vitros 950 analyzer (Johnson & Johnson Clinical Diagnostics Inc., Rochester, NY). Creatinine levels were calibrated to the Cleveland Clinic standard (0.9954*Cr + 0.0208)^16^. CRP was measured using a BNII nephelometer (N high sensitivity CRP; Dade Behring Inc.), with intra-assay CVs of 2.3% – 4.4%, inter-assay CVs of 2.1% – 5.7%, and a detection level of 0.18 mg/L^17^. DNA was isolated from peripheral leukocytes in the Buffy coat using the Gentra Puregene Blood Kit. CRP and complete blood counts measurements from MESA Exam 5 (2010-2012) were analyzed in a similar manner.

The primary endpoint was incidence of all-cause mortality, which was verified in 12-month intervals by a telephone interview with participants, reviewing copies of death certificates and medical records for all hospitalizations, and annual National Death Index queries. The secondary endpoint of incident heart disease (which was defined as composite of coronary heart disease mortality and non-fatal myocardial infarction in JHS and coronary heart disease mortality, and resuscitated cardiac arrest, and non-fatal myocardial infarction in MESA) and heart failure were verified in a similar manner. In JHS, surveillance of mortality and incident coronary disease spanned from 2005 to 2016, whereas surveillance of heart failure events spanned from 2005 to 2016. In MESA, surveillance period of all outcomes spanned from 2001 to 2018.

Continuous and categorical variables are presented as mean with one standard deviation and were compared with one-way t-test or ANOVA for means and chi-square test for proportions. Linear regression models were applied to determine the independent variables associated with serum hs-CRP levels. The following demographic and clinical covariates were included in the model: age, sex, BMI, history of hypertension, fasting glucose, serum creatinine, LDL-C, percent African ancestry, Duffy receptor status, and study cohort. The Duffy gene and hs-CRP gene are in relative proximity on chromosome 1. To account for the possibility of long-range linkage disequilibrium, sensitivity analysis with a continuous linear regression model including the Duffy null variable was performed while simultaneously accounting for eight *CRP* locus SNP alleles known to influence circulating levels of CRP: rs11265259, rs1800947, rs2211321, rs7551731, rs73024795, rs553202904, rs12734907, rs181704186^18^.

Event rates were estimated using the Kaplan-Meier time-to-event methodology and compared using log-rank tests. Multivariable Cox proportional hazard regression was used to determine the independent predictors of the primary and secondary outcomes. The following demographic and clinical covariates were simultaneously included in the model for the primary and secondary outcomes: age, sex, BMI, history of hypertension, fasting plasma glucose, serum creatinine, LDL-C, Duffy status, and percent African ancestry.

The significance level for all analyses was set at p<0.05 (two-sided). All analyses were performed with IBM SPSS Statistics for Mac, version 24 (IBM Corp., Armonk, N.Y., USA) and GraphPad Prism 9 version 9.2.0 (283) for Mac, GraphPad Software, La Jolla California USA (www.graphpad.com).

## Results

From the JHS and MESA cohorts, 4,307 participants (2,942 Duffy null and 1,365 Duffy receptor positive) were included in this analysis. Baseline characteristics, by Duffy receptor status, are displayed in Table 1. Participants in the Duffy null and positive groups were well-balanced except for differences in the percent of their genomes of African descent, younger age, levels of triglycerides and hs-CRP.

**Table 1.** Baseline characteristics by Duffy antigen receptor status.

|  | Duffy antigen<br>receptor negative<br>(n=2,942) | Duffy antigen<br>receptor positive<br>(n=1,365) | p-value |
| --- | --- | --- | --- |
| **Demographics** |  |  |  |
| Age (years) | 57 ± 12 | 58 ± 12 | <0.001 |
| Males (%) | 1151 (39) | 543 (39) | n.s. |
| Percent African<br>ancestry | 86 ± 0.08 | 77 ± 0.13 | <0.001 |
| **Clinical variables** |  |  |  |
| Hypertension (%) | 1667 (57) | 768 (56) | n.s. |
| BMI (kg/m2) | 30.4 ± 6.9 | 30.1 ± 7.0 | n.s. |
| Fasting plasma<br>glucose (mg/dL) | 92 ± 33.6 | 93 ± 30.4 | n.s. |
| Total cholesterol<br>(mg/dL) | 195 ± 40.6 | 194 ± 38.5 | n.s. |
| Triglycerides<br>(mg/dL) | 89 ± 81.9 | 93 ± 68.6 | 0.01 |
| HDL cholesterol<br>(mg/dL) | 50 ± 14.8 | 50 ± 14.6 | n.s. |
| LDL cholesterol<br>(mg/dL) | 122 ± 36.8 | 122 ± 35.6 | n.s. |
| Serum creatinine<br>(mg/dL) | 0.94 ± 0.53 | 1.02 ± 0.43 | n.s. |
| hs-CRP (mg/L) | 2.7 ± 10.1 | 2.1 ± 7.00 | <0.001 |

BMI – body mass index; HDL – high-density lipoprotein; hs-CRP – high-sensitivity c-reactive protein; LDL – low-density lipoprotein; LVH – left ventricular hypertrophy

Linear regression modeling identified male gender (negative effect), BMI, fasting glucose, LDL-C (negative effect), and Duffy status as independent variables associated with serum hs-CRP levels. Age, hypertension, serum creatinine, study cohort, and percent African ancestry were not associated with hs-CRP levels (Table 2a).

**Table 2a.**
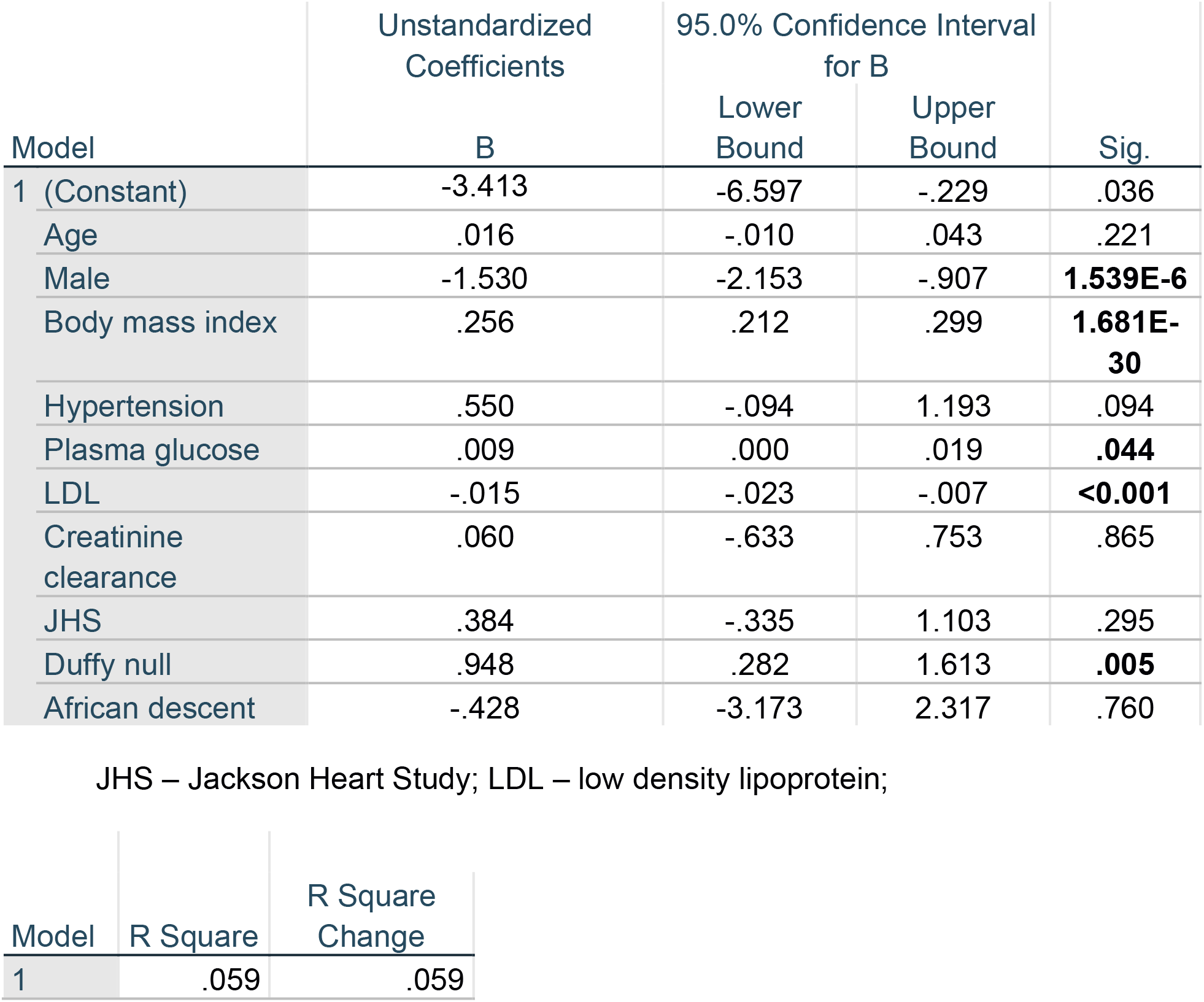
Independent predictors of hs-CRP.

| Model | Unstandardized Coefficients<br>B | 95.0% Confidence Interval<br>for B |  | Sig. |
| --- | --- | --- | --- | --- |
|  |  | Lower Bound | Upper Bound |  |
| 1 (Constant) | -3.413 | -6.597 | -.229 | .036 |
| Age | .016 | -.010 | .043 | .221 |
| Male | -1.530 | -2.153 | -.907 | **1.539E-6** |
| Body mass index | .256 | .212 | .299 | **1.681E-30** |
| Hypertension | .550 | -.094 | 1.193 | .094 |
| Plasma glucose | .009 | .000 | .019 | **.044** |
| LDL | -.015 | -.023 | -.007 | **<0.001** |
| Creatinine clearance | .060 | -.633 | .753 | .865 |
| JHS | .384 | -.335 | 1.103 | .295 |
| Duffy null | .948 | .282 | 1.613 | **.005** |
| African descent | -.428 | -3.173 | 2.317 | .760 |

JHS – Jackson Heart Study; LDL – low density lipoprotein;
| Model | R Square | R Square Change |
| --- | --- | --- |
| 1 | .059 | .059 |

Linear regression models also did not find an effect of the Duffy null genotype on hs-CRP levels once conditioning for distinct *CRP* locus alleles in linkage disequilibrium with the Duffy gene (Table 2b)^18^.

**Table 2b.** Linear regression models of Duffy null and CRP alleles in continuous model.

| Coefficient | Estimate | Lo 95%CI | Hi 95%CI | P-val |
| --- | --- | --- | --- | --- |
| (Intercept) | 5.5667 | 4.6279422 | 6.5053618 | 2.00E-16 |
| Duffy.null | 0.3855 | -0.3473321 | 1.1183374 | 0.302587 |
| rs11265259 | -1.3616 | -2.1143912 | -0.6088001 | 0.000397 |
| rs1800947 | -1.3904 | -3.5363776 | 0.7555743 | 0.204195 |
| rs2211321 | -1.0546 | -1.653387 | -0.4557823 | 0.000562 |
| rs7551731 | -0.9199 | -1.4857942 | -0.3540476 | 0.001452 |
| rs73024795 | 2.1242 | 1.4459884 | 2.8024926 | 9.08E-10 |
| rs553202904 | -2.4508 | -11.4793492 | 6.5777663 | 0.594733 |
| rs12734907 | -0.4768 | -1.4140775 | 0.460387 | 0.318728 |
| rs181704186 | -2.107 | -4.1106036 | -0.1033385 | 0.039358 |

The primary endpoint of all-cause mortality (n=939, 24.5%) occurred in 25.6% with Duffy null and 22.1% with Duffy receptor positive participants (p=n.s., Figure 1a). Cox regression modeling of all-cause mortality identified age, male sex, hypertension, plasma glucose, hs-CRP, creatinine clearance, and Duffy null status as independent variables associated with all-cause mortality (Table 3a). Duffy receptor status was not a significant predictor of incident heart failure (Table 3b; 7.2% Duffy null and 7.6% Duffy receptor positive, also Figure 1b). Similarly, Duffy receptor status was not a significant predictor of incident coronary heart disease (Table 3c; 4.7% Duffy receptor status negative and 5.1% Duffy receptor status positive, also Figure 1c).

**Figure 1.**
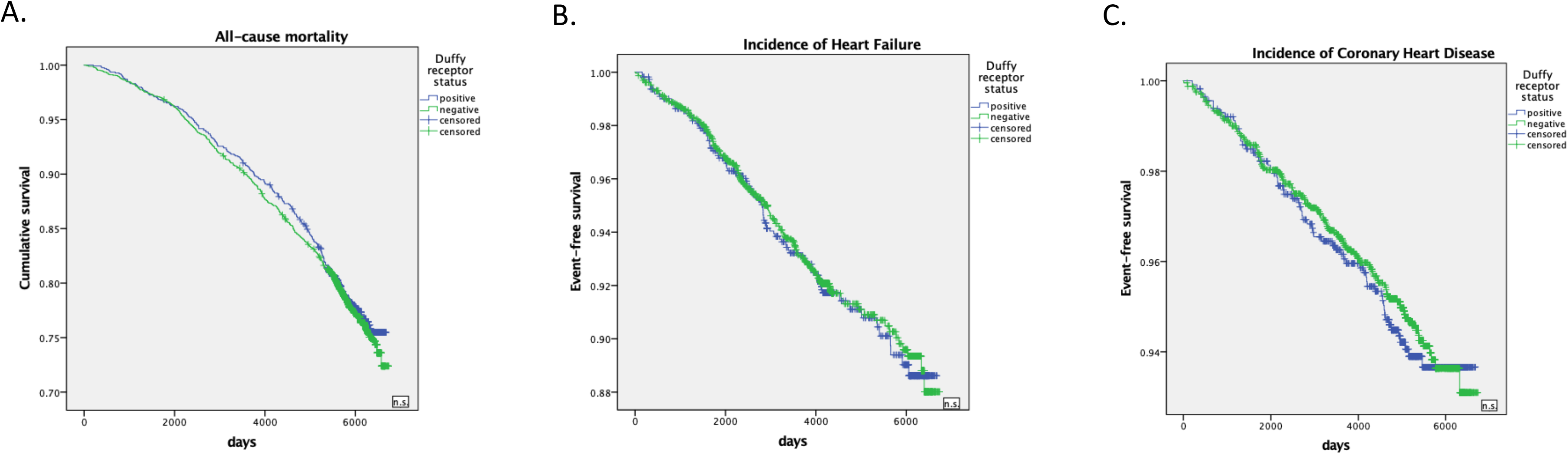
Kaplan Meier curves for primary and secondary outcomes.

**Table 3a.** Cox regression model for all-cause mortality.

**Model 1**
|  | Hazard Ratio | 95.0% CI for Hazard Ratio |  | p value |
| --- | --- | --- | --- | --- |
|  |  | Lower | Upper |  |
| Age | 1.087 | 1.079 | 1.095 | **1.177E-112** |
| Male | 1.351 | 1.175 | 1.553 | **2.328E-5** |
| BMI (per kg/m2) | 1.006 | .994 | 1.017 | .314 |
| Hypertension | 1.509 | 1.281 | 1.778 | **9.012E-7** |
| Plasma glucose | 1.005 | 1.003 | 1.007 | **5.024E-9** |
| LDL | 1.000 | .998 | 1.002 | .765 |
| Hs-CRP (per mg/L) | 1.012 | 1.007 | 1.018 | **1.750E-5** |
| Creatinine clearance | 1.429 | 1.316 | 1.553 | **3.132E-17** |
| Duffy null status | .839 | .727 | .969 | **.017** |

**Table 3b.** Cox regression model for incident heart failure.

|  | **Hazard Ratio (95% CI)** | **p-value** |
| --- | --- | --- |
| Age (per year) | 1.07 (1.05, 1.08) | <0.001 |
| Male | 1.35 (1.04, 1.71) | 0.02 |
| BMI (per kg/m<sup>2</sup>) | 1.04 (1.02, 1.06) | <0.001 |
| Hypertension | 1.78 (1.32, 2.40) | <0.001 |
| Fasting plasma glucose<br>(per mg/dL) | 1.007 (1.005, 1.01) | <0.001 |
| serum creatinine (per<br>mg/dL) | 1.39 (1.21, 1.59) | <0.001 |
| LDL cholesterol | 1.001 (0.99, 1.005) | n.s. |
| hs-CRP (per mg/L) | 1.004 (0.99, 1.02) | n.s. |
| Percent African<br>ancestry | 2.02 (0.767, 6.14) | n.s. |
| Duffy antigen receptor<br>genotype negative | 1.05 (0.79, 1.39) | n.s. |

BMI – body mass index; hs-CRP – high-sensitivity c-reactive protein; LDL – low-density lipoprotein

**Table 3c.** Cox regression model for incident coronary heart disease.

|  | **Hazard Ratio (95% CI)** | **p-value** |
| --- | --- | --- |
| Age (per year) | 1.05 (1.04, 1.07) | <0.001 |
| Male | 1.78 (1.31, 2.41) | <0.001 |
| BMI (per kg/m<sup>2</sup>) | 0.99 (0.96, 1.02) | n.s. |
| Hypertension | 1.93 (1.34, 2.79) | <0.001 |
| Fasting plasma glucose<br>(per mg/dL) | 1.006 (1.002, 1.009) | 0.002 |
| serum creatinine (per<br>mg/dL) | 1.38 (1.16, 1.65) | <0.001 |
| LDL cholesterol | 1.003 (0.99, 1.007) | n.s. |
| hs-CRP (per mg/L) | 1.01 (1.004, 1.02) | 0.005 |
| Percent African<br>ancestry | 0.40 (0.15, 1.98) | n.s. |
| Duffy antigen receptor<br>negative | 1.21 (0.85, 1.72) | n.s. |

BMI – body mass index; hs-CRP – high-sensitivity c-reactive protein; LDL – low-density lipoprotein

## Discussion

The major findings from this analysis of Black participants of the JHS and MESA are as follows: 1) null mutations of the Duffy receptor gene was not independently associated with elevated levels of serum hs-CRP levels once conditioning for *CRP* locus alleles known to be associated with changes in the expression level of CRP 2) There was no difference in adverse outcomes based on Duffy null status over the follow-up period.

Our findings are novel as it is the first to study the inflammatory cytokine hs-CRP in relation to Duffy receptor status in a systematic manner and describe its potential effects on long-term cardiovascular outcomes. Hs-CRP levels was not statistically significantly higher in those with null mutations in the Duffy receptor gene once conditioning for the 8 *CRP* locus alleles that may be in linkage disequilibrium with the Duffy gene. These 8 variants were identified using whole genome sequencing studies which have cataloged loci and alleles associated with differencing levels of hs-CRP in TOPMed cohorts, including JHS and MESA^18^, but did not identify the Duffy allele as a genome-wide significant signal as it was located within the CRP region (chr1:158671755-159791734) (p=2.8×10-11 in pooled ancestry analysis). Although that study did not stratify analyses by sex and utilized additive models which may not be appropriate for studying SNP phenotypes of the Duffy gene that are recessive in nature it did not reveal SNP rs2814778 with genome-wide significance while conditioning on all 8 CRP variants (p=0.0002)^18^. A separate protein expression study demonstrated statistically significant levels of hs-CRP levels based on SNP rs2814778 genotype at a p<0.015, however this was a secondary analysis and did not account for the associated CRP locus^8^. This study confirms that the association of CRP levels with the SNP rs2814778 is largely dependent on CRP locus alleles known to influence levels of CRP in serum and in linkage disequilibrium to the Duffy gene ^8,19^.

Overall, in the MESA cohort, Black participants have higher incidence of all-cause mortality^1^, which is not explained by Duffy receptor status. Duffy receptor status was no longer a statistically significant variable of all-cause mortality after consideration of percent African descent demonstrating that the Duffy null genotype is an ancestry informative marker in Black adults. Thus, this indicates any possible effect by Duffy receptor status on mortality in Black individuals is not significant^20^.

## Limitations

We recognize important limitations to our study design, observations, and conclusions. The first is the relatively healthier cohort of participants with no clinical cardiovascular disease at baseline enrolled in MESA, which may not be generalizable to the entire US population. Second, only baseline demographic and risk factors were considered in our analysis, thus it is unclear whether certain risk factors progressed or recessed over time in study participants.

## Conclusions

Hs-CRP was significantly higher in the Duffy null group; however, this was dependent on linkage disequilibrium of the CRP gene in proximity of the Duffy gene. Duffy null status was not found to be independently associated with higher incidence of all-cause mortality after adjusting for possible confounders in Black adults. Given the ongoing health inequities in cardiovascular outcomes in the Black population more needs to be done to understand both the determinants driving such outcomes so that personalized therapies and risk modification schema may be developed and implemented to addresses these disparities.

## Data Availability

available upon approval from JHS and MESA

## Acknowledgement/Disclaimer

The Jackson Heart Study (JHS) is supported and conducted in collaboration with Jackson State University (HHSN268201800013I), Tougaloo College (HHSN268201800014I), the Mississippi State Department of Health (HHSN268201800015I) and the University of Mississippi Medical Center (HHSN268201800010I, HHSN268201800011I and HHSN268201800012I) contracts from the National Heart, Lung, and Blood Institute (NHLBI) and the National Institute on Minority Health and Health Disparities (NIMHD). The authors also wish to thank the staffs and participants of the JHS.

Whole genome sequencing (WGS) for the Trans-Omics in Precision Medicine (TOPMed) program was supported by the National Heart, Lung and Blood Institute (NHLBI). WGS for “NHLBI TOPMed: Multi-Ethnic Study of Atherosclerosis (MESA)” (phs001416.v1.p1) was performed at the Broad Institute of MIT and Harvard (3U54HG003067-13S1). Centralized read mapping and genotype calling, along with variant quality metrics and filtering were provided by the TOPMed Informatics Research Center (3R01HL-117626-02S1). Phenotype harmonization, data management, sample-identity QC, and general study coordination, were provided by the TOPMed Data Coordinating Center (3R01HL-120393-02S1), and TOPMed MESA Multi-Omics (HHSN2682015000031/HSN26800004). The MESA projects are conducted and supported by the National Heart, Lung, and Blood Institute (NHLBI) in collaboration with MESA investigators. Support for the Multi-Ethnic Study of Atherosclerosis (MESA) projects are conducted and supported by the National Heart, Lung, and Blood Institute (NHLBI) in collaboration with MESA investigators. Support for MESA is provided by contracts 75N92020D00001, HHSN268201500003I, N01-HC-95159, 75N92020D00005, N01-HC-95160, 75N92020D00002, N01-HC-95161, 75N92020D00003, N01-HC-95162, 75N92020D00006, N01-HC-95163, 75N92020D00004, N01-HC-95164, 75N92020D00007, N01-HC-95165, N01-HC-95166, N01-HC-95167, N01-HC-95168, N01-HC-95169, UL1-TR-000040, UL1-TR-001079, UL1-TR-001420, UL1TR001881, DK063491, and R01HL105756. The authors thank the other investigators, the staff, and the participants of the MESA study for their valuable contributions. A fill list of participating MESA investigators and institutes can be found at http://www.mesa-nhlbi.org.

*The views expressed in this manuscript are those of the authors and do not necessarily represent the views of the National Heart, Lung, and Blood Institute; the National Institutes of Health; or the U.S. Department of Health and Human Services*.

